# Gulf War veterans exhibit broadband sleep EEG power reductions in regions overlying the frontal lobe

**DOI:** 10.1101/2021.04.26.21251831

**Authors:** Eric W. Moffet, Stephanie G. Jones, Theodore Snyder, Brady Riedner, Timothy Juergens

## Abstract

**Aims:** Nearly a third of U.S. veterans who deployed in support of the 1990-1991 Persian Gulf War are affected by Gulf War illness (GWI). Here we aimed to characterize whether subjective sleep complaints in GWI veterans are associated with objective sleep EEG disturbances relative to healthy veterans and controls; and whether Gulf War veterans show alterations in neural activity during sleep that differentiate them from healthy subjects.

**Main methods:** We used high-density EEG (HDEEG) to assess regional patterns of rapid eye movement (REM) sleep and non-REM (NREM) sleep between three groups: Gulf War male veterans with fatigue and GWI, Gulf War male veterans without fatigue or GWI, and control males. The groups were matched relative to age, sex and obstructive sleep apnea. Topographic comparisons of nocturnal NREM and REM sleep were made between groups for all frequency bands.

**Key findings:** Topographic analysis revealed a broadband reduction in EEG power in a circumscribed region overlying the frontal lobe in both groups of Gulf War veterans, regardless of GWI and fatigue. This frontal reduction in neural activity was present, to some extent, across all frequency bands in NREM and REM sleep.

**Significance:** Given that our findings were observed in all Gulf War veterans, it appears unlikely that frontal sleep HDEEG power reductions prove wholly responsible for fatigue symptoms. These results provide avenues for research and underpin the importance of maintaining a high index of suspicion when providing clinical care to formerly deployed veterans of the Persian Gulf War.

## INTRODUCTION

Gulf War illness (GWI) affects up to one third of the 697,000 U.S. veterans of the 1990-1991 Persian Gulf War (1, 2). Although clinical presentations prove heterogeneous, prominent symptoms include fatigue, sleep disturbances, musculoskeletal pain, and alterations in mood and cognition (1). The precise mechanisms involved in the development of GWI remain unclear. However, the Gulf War was remarkable for the extent of chemical-exposures experienced by troops including an array of known neurotoxins from chemical warfare agents, pesticides, and prophylactic medications. Given this exposure profile and given that the most prominent symptoms of GWI involve nervous system dysfunction, etiological research has focused predominantly on the central nervous system (1-5).

Although subjective sleep complaints are a central feature of the GWI phenotype —and sleep disturbance has notable effects on the other hallmark symptoms of pain, fatigue, mood, and cognitive performance—remarkably little research has focused on characterizing sleeping brain function in this population. Though the basic sleep architecture of veterans with GWI has appeared normal in previous studies (1-3), the frontal lobe is thought to be vital to the restorative function of sleep (6, 7), and a battery of literature demonstrates frontal lobe abnormalities in GWI—including deficits in function, performance, and perfusion, as well as pathologies of the gray matter (GM) and white matter (WM) (8-19). In light of these relationships, it is conceivable that sleep pathology may play a significant role in the maintenance of the disorder. An equally plausible scenario is that sleep pathology, along with other CNS symptoms common to GWI (20,21), arise as a secondary consequence of neurotoxin exposure. However, to date, a paucity of electrophysiological data exists to characterize brain activity during sleep in this population, and it is not known whether or to what extent objective features of sleep may be adversely affected.

Previous GWI sleep studies used polysomnography with a 10-20 EEG montage (22-24). This montage possesses limited ability to detect spontaneous neural activity. In contrast, high - density EEG (HDEEG; 256-electrodes) affords robust spatiotemporal localizing capacity (25). Sleep HDEEG shows reproducibility between subjects across multiple nights, while minimizing the artefactual confounds of wake (26, 27). Our group has used this technology to explore the pathophysiology of insomnia, night terrors, sleep apnea, stroke, and epilepsy (28-32).

The brain physiology of veterans with GWI remains in question. In this study, we used HDEEG to record neural activity during sleep in Gulf War veterans with GWI who endorse significant fatigue (GWIV), Gulf War veterans without GWI (HGWV), and healthy civilian control subjects (CC) to determine (1) whether subjective sleep complaints in veterans with GWI are associated with objective sleep HDEEG disturbances relative to healthy veterans and healthy controls; and (2) whether Gulf War veterans show alterations in neural activity during sleep that differentiate them from healthy subjects.

## MATERIALS AND METHODS

### Subjects

This prospective, two arm, case-control study considers military veterans that deployed in support of the 1990-1991 Persian Gulf War. We consider both healthy Gulf War veterans (HGWV) and veterans who specifically report fatigue in the context of GWI (i.e., GWIV). A GWI diagnosis was confirmed using CDC-defined chronic multisymptom illness criteria, described as having one or more symptoms from at least two separate clusters (fatigability, mood and cognition, musculoskeletal) for six months or more (33, 34). We also consider a cohort of healthy civilian controls (CC), selected from a separate study. Groups were screened for the absence of neurologic, sleep, and psychiatric disorders in addition to use of medications indicated below. All three groups were comparable in age, sex, and obstructive sleep apnea status.

Veterans were recruited from the Madison, WI, William S. Middleton Memorial Veterans Hospital. The study was conducted at Wisconsin Sleep. Subjects were recruited via flier endorsed by the U.S. Department of Veterans Affairs and an informational letter sent to 800+ veterans on the William S. Middleton Memorial Veterans Hospital’s Persian Gulf War registry. Interested participants were first evaluated with the Fatigue Severity Scale (FSS; expand description provided below). Literature indicates that a minimum score of four indicates pathologic fatigue (35). GWIV were included when their FSS score was at least five with a duration of fatigue greater than one month; HGWV and control groups were limited to a maximum FSS score of two. Next, eligible participants were assessed via the Structured Clinical Interview for DSM-IV disorders, which is a structured psychiatric interview used to identify common DSM-IV diagnoses. Those with major depressive disorder, generalized anxiety disorder, substance abuse or dependence, schizophrenia, bipolar disorder, post-traumatic stress disorder, or obsessive-compulsive disorder were excluded. Subjects were also excluded if they had a diagnosis of restless leg syndrome or periodic limb movements of sleep. Candidates with body mass index >40 kg/m2 were also excluded. Obstructive sleep apnea (OSA) patients were excluded. Subjects that displayed evidence of OSA during the study were not excluded; this finding was tracked and accounted for in the analyses. Participants were screened out for use, within 2 weeks of the study, of sleep medications (trazodone, zolpidem, zolpidem, ramelteon, eszopiclone, zaleplon, melatonin, estazolam, temazepam), sedating medications (tricyclic antidepressants, mirtazapine, benzodiazepine, opiates, anticonvulsants, beta blockers), and/or stimulating medications (bupropion, modafinil, amphetamine, dextroamphetamine, steroids). All subjects were instructed to maintain normal sleep-wake cycles the week prior to study participation; in order to screen out subjects with dysregulated sleep-wake cycles, participants kept a sleep diary and underwent actigraphy during this time to insure they spent 7-9 hours per night in bed on average with a bedtime between 21:00-00:00 and wake time between 05:00-09:00.

Each participant provided informed consent for experimentation with human subjects. The Institutional Review Board at the University of Wisconsin-Madison approved the study.

### Symptom Variable Surveys

Subjective fatigue and sleepiness were assessed using the FSS and the Insomnia Severity Index (36, 37). The FSS is a self-administered questionnaire with 9 items (questions) investigating the severity of fatigue in different situations during the past week. Scoring of each item ranges from one to seven, where one indicates strong disagreement and seven strong agreement. The final score represents the mean value of the nine items. ISI was used to assess self-reported insomnia severity. The ISI consists of seven items with responses ranked from zero to four, producing total scores of zero to twenty-eight. Total ISI scores can be categorized into the following: no clinically significant insomnia (zero to seven), subthreshold insomnia mild (eight to fourteen), moderate clinical insomnia (fifteen to twenty-one), and severe clinical insomnia (twenty-two to twenty-eight).

### HDEEG Polysomnography

Subjects underwent overnight HDEEG polysomnography utilizing 256-electrode EEG nets (Electrical Geodesics, Inc. (EGI), Eugene, OR). Subjects were monitored with standard electrocardiogram, submental and bilateral tibial electromyogram, electrooculogram, pulse oximetry, a position sensor, and respiratory inductance plethysmography. Participants arrived at Wisconsin Sleep by 18:00 hours, allowing for setup, and went to bed at their normal time, but no later than 00:00 hours. Participants were allowed to sleep undisturbed in the lab until no later than 06:30 hours. A registered sleep technologist oversaw the night and conducted initial sleep scoring; a certified sleep medicine physician approved each report. Alice® Sleepware (Philips Respironics, Murrysville, PA) and AASM guidelines were utilized for sleep staging, based on six mastoid re-referenced HDEEG channels approximately located at 10-20 EEG locations (F3, F4, C3, C4, O1, O2).

### HDEEG Recordings

All-night recordings of sleep using HDEEG were sampled at 500 Hz (vertex referencing) using NetStation software (EGI) and a NetAmps 300 amplifier. Initially, a 0.1 Hz first-order high-pass filter was applied, which eliminates low frequency drift, mimicking common analog filters. Band-pass filtering was then completed on the data (Kaiser type, 0.3 - 40 Hz), followed by downsampling to 128 Hz. The data was high-pass filtered once more, (two-way least-squares Frequency Infinite Response Filter, 1 Hz) to remove sweat artifact. Bad channels were manually removed, after visual inspection for scalp contact interruption and high-frequency noise. Then, using MATLAB (The MathWorks, Inc., Natick, MA), the data was interpolated, and average referenced. Independent component analysis was conducted and cardiac, eye movement, and muscle artifact components removed. Spectral analysis was completed in six second consecutive epochs (Welch’s method with Hamming window). Finally, spectral density was computed for six frequencies: delta, 1–4 Hz; theta, 4–8 Hz; alpha, 8–12 Hz; sigma, 12–16 Hz; beta, 16–25 Hz; and gamma, 25-40 Hz.

### Statistics

Statistical analyses were conducted with Microsoft Excel and MATLAB. Sleep architecture, demographic data, and global spectral EEG data were examined for differences between groups using a one-way ANOVA F test (alpha = 0.05) when three groups were considered and unpaired t-tests when two groups were considered. Unpaired *t*-tests were conducted on electrode-to-electrode topographic NREM and REM data. Statistical nonparametric mapping, with suprathreshold cluster testing, was utilized to identify significant electrode clusters; this approach provides a thorough assessment of significance for absolute and normalized topographic power maps (38). In practice, it addresses the multiple comparisons problem and applies weak distributional assumptions. It is thus applicable for small sample sizes where normality of data proves difficult to define. Briefly, a t-value threshold was first selected (t-value = 2 was chosen across all frequencies), then topographic power maps were shuffled randomly between groups in all possible combinations. The maximum cluster above threshold of each reshuffling was used to define a maximum cluster size distribution. By comparing actual cluster size to the maximal cluster distribution, a suprathreshold cluster p-value was established. In order to account for interindividual variability in the absolute data, we normalized topographic maps with z-scoring of all channels across all subjects. Signal-to-noise ratios were increased via statistical analyses of topographic data that excluded electrodes on the neck and face (i.e., using EEGLAB software, a plugin for MATLAB; the “topoplot” function was employed to specify electrodes within a 0.57 plotting radius); as a result, 173 channels layered the scalp.

To assess relationships between sleep abnormalities and subjective complaints of fatigue and sleep disturbance in GWI, scores on the FSS and ISI were correlated with normalized EEG SWA power in the cluster identified as significantly different between groups using Pearson’s R.

### Data Availability

The authors will make available, upon reasonable request, the raw data utilized for this manuscript. Inquiries should be directed to the corresponding author. Requests must comply with the University of Wisconsin’s ethical approvals and data-sharing policies.

## RESULTS

### Demographic Data

**Table 1** summarizes participant information and sleep architecture variables. The study contained three groups of nine males with physiologically comparable ages. CC and GWIV possessed mean ages of 42.3 (± 1.0) and 41.8 (± 0.6) years, respectively; the average age of HGWV was 44.6 (± 1.2) years. The ANOVA for age revealed a significant difference across groups, F(2,24) = 3.84, p = 0.036. Follow-up t-tests revealed this difference to lie between the HGWV and CC groups (HGWV mean 44.56 (+/-1.2) years, CC mean 41.22 (+/-0.8) years, p = .049, Bonferroni). Both HGWV and CC groups were statistically similar in age to the GWIV group (p = 1 and p = .124, Bonferroni). Sleep architecture, including arousal index, total sleep time, wake after sleep onset, sleep efficiency, sleep latency, REM latency, and percentage of time in each sleep stage compared to overall sleep, did not vary among groups. Crucially, the presence of OSA, as represented by the mean arousal index, also did not differ across groups (F(2,24) = 0.23, p = 0.80).

**Table 1.** Demographics, sleep characteristics, and symptom variables for GWIV, CC, and HGWV groups. Mean (standard error) is presented for each group, with p-values from ANOVA when data available for three groups, and from unpaired t-test when data available for Gulf War groups only. An * indicates a significant difference among groups (p < .05). AHI = apnea-hypopnea index; AI = arousal index; CC = civilian control; FSS = Fatigue Severity Scale; FOSQ = Functional Outcomes of Sleep Questionnaire; GWIV = Gulf War illness veteran; HGWV = healthy Gulf War veteran; ISI = Insomnia Severity Index; MFSI = Multidimensional Fatigue Symptom Inventory; N1% = total stage N1 sleep per TST; N3% = total stage N1 sleep per TST; N3% = total stage N3 sleep per TST; REM% = total rapid eye movement (REM) sleep per TST; REML = REM sleep latency; SE = sleep efficiency; SL = sleep latency; WASO = wake after sleep onset; TST = total sleep time

|  | GWIV | CC | HGWV | p |
| --- | --- | --- | --- | --- |
| Age* | 41.78 (0.6) | 41.22 (0.8) | 44.56 (1.2) | 0.036 |
| AHI | 4.00 (1.9) | 3.77 (2.0) | 5.00 (2.0) | 0.929 |
| AI | 17.73 (5.6) | 19.5 (2.3) | 22.23 (5.5) | 0.795 |

**Table 1. Demographics, sleep characteristics, and symptom variables for GWIV, CC, and HGWV groups.**
|  |  |  |  |  |
| --- | --- | --- | --- | --- |
| TST | 364.78 (46.2) | 343.67 (21.2) | 354.39 (11.9) | 0.885 |
| WASO | 86.83 (26.0) | 89.39 (18.4) | 90.61 (11.9) | 0.990 |
| SE | 70.32 (5.3) | 72.82 (3.8) | 74.71 (2.9) | 0.753 |
| SL | 35.11 (16.0) | 16.89 (2.8) | 13.33 (3.8) | 0.248 |
| REML | 130.44 (24.7) | 145.06 (27.1) | 157.56 (18.8) | 0.724 |
| N1% | 13.79 (4.2) | 9.81 (1.3) | 11.33 (1.9) | 0.596 |
| N2% | 60.82 (4.9) | 63.84 (1.5) | 60.75 (2.0) | 0.735 |
| N3% | 13.22 (4.1) | 11.75 (2.5) | 12.62 (2.6) | 0.946 |
| REM% | 12.20 (2.9) | 14.68 (1.9) | 16.16 (1.6) | 0.451 |
| FSS* | 5.52 (0.5) | 2.32 (0.2) | 2.27 (0.2) | <0.001 |
| ISI* | 17.67 (2.4) | 2.89 (0.9) | 5.22 (1.5) | <0.001 |

Significant differences were observed among group scores on the FSS F(2,24) = 31.16, p <.001 and ISI F(2,24) = 21.76, p < .001. Follow up analysis on the FSS showed the GWIV group scored significantly higher than the CC group (GWIV mean 5.52 (+/ −0.5), CC mean 2.32 (+/- 0.2), p < .001, Bonferroni) and the HGWV group (GWIV mean 5.52 (+/-0.5), HGWV mean 2.27 (+/-0.2), p < .001). Similarly, on the ISI, the GWIV group scored higher than the CC group (GWIV mean 17.67 (+/-2.4), CC mean 2.89 (+/-0.9), p < .001, Bonferroni) and HGWV group (GWIV mean 17.67 (+/-2.4), HGWV mean 5.22 (+/-1.5), p < .001, Bonferroni). Box plots of ISI and FSS scores are presented in **Figure 1**, panels A and B, respectively .

**Figure 1.**
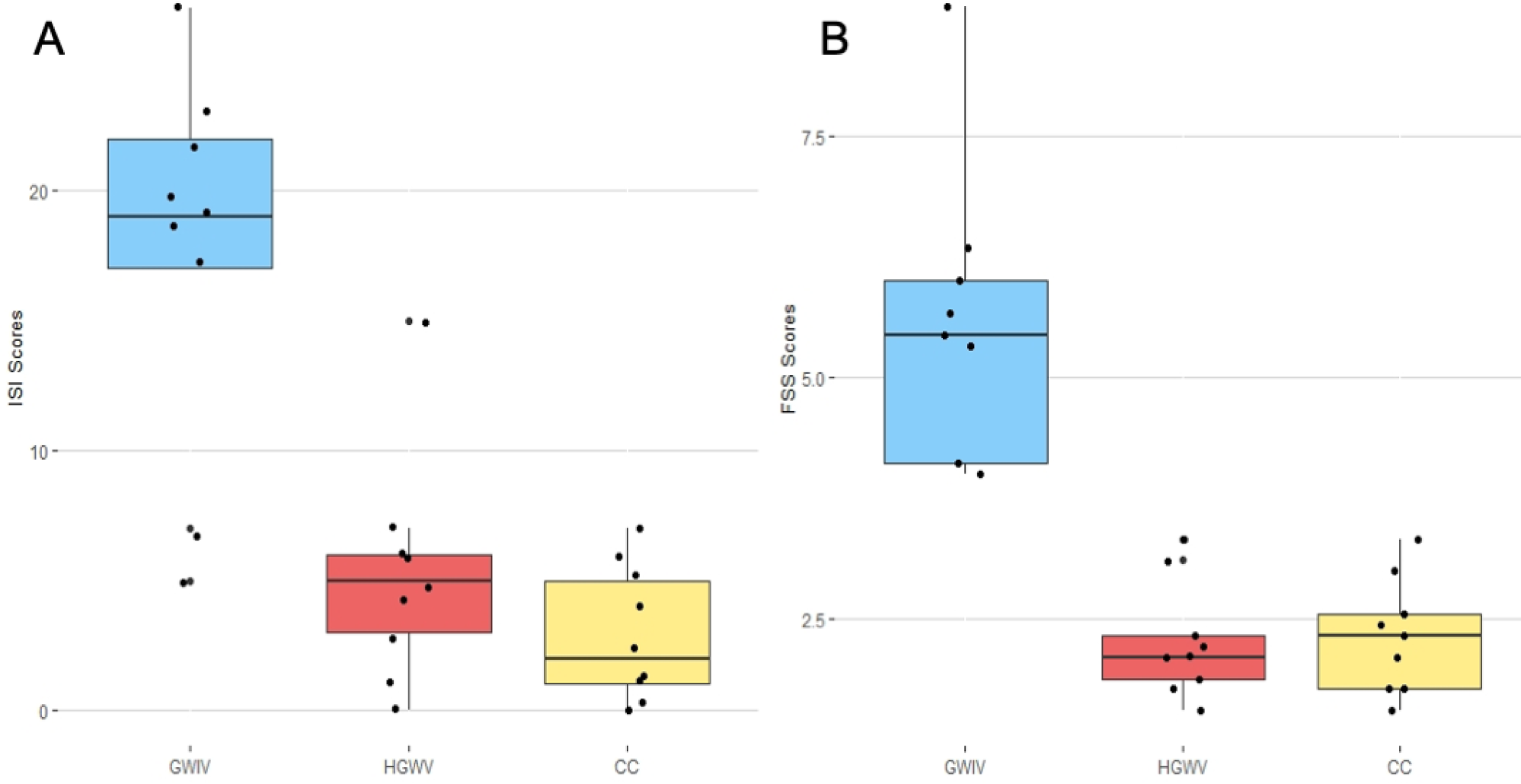
Box plots of ISI (A) and FSS (B) scores by group. Median scores for each group (blue = GWIV, red = HGWV, yellow = CC) are represented by the line in the middle of each box, and the box edges represent the 1st and 3rd quantiles of the data for each group. Lines extend to the lowest and highest values in each group that are not considered outliers. Black dots represent data points. A one-way ANOVA showed that ISI scores (A) differed significantly according to group F(2,24) = 21.76, p < .001, and so did FSS scores (B) F(2,24) = 31.16, p <.001. Follow up analysis showed the GWIV group scored higher than CC (GWIV mean 17.67 (+/-2.4), CC mean 2.89 (+/-0.9), p < .001, Bonferroni) and HGWV (GWIV mean 17.67 (+/-2.4), HGWV mean 5.22 (+/-1.5), p < .001, Bonferroni) on the ISS (A) and higher than CC (GWIV mean 5.52 (+/-0.5), CC mean 2.32 (+/-0.2), p < .001, Bonferroni) and HGWV (GWIV mean 17.67 (+/-2.4), HGWV mean 5.22 (+/-1.5), p < .001, Bonferroni) on the FSS (B). CC and HGWV groups did not significantly differ on either measure. CC = civilian control; GWIV = Gulf War illness veteran; HGWV = healthy Gulf War veteran

### HDEEG Polysomnography Data

We examined how global sleep power varied between groups by comparing spectral data averaged across channels. **Figure 2** illustrates spectral profiles for CC, HGWV, and GWIV groups within NREM (panel A) and REM (panel B) sleep and the associated p-values for ANOVA tests for group differences in each 1/6 Hz frequency bin, from 1/6 – 40 Hz. All three groups displayed statistically similar distributions of global EEG power in NREM sleep (ANOVA p > .05).

**Figure 2.**
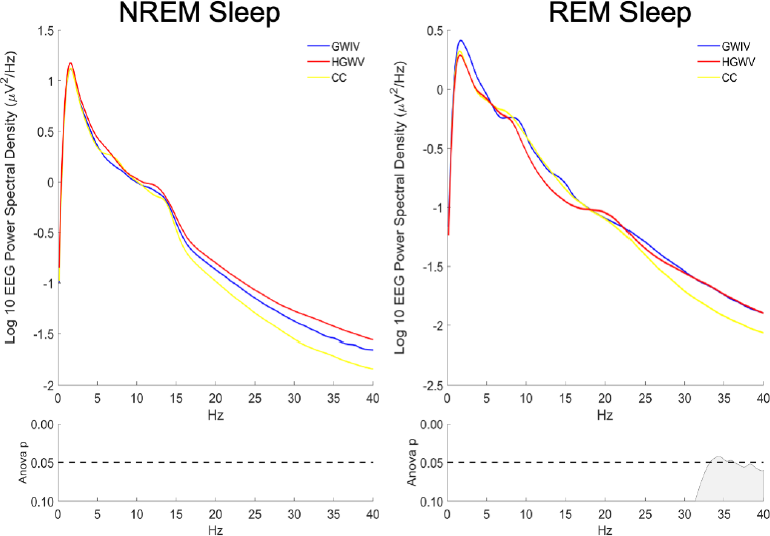
GWIV, HGWV, and CC exhibit similar global spectral profiles in all-night NREM sleep, CC show lower gamma in REM. Mean log10 EEG power (y-axis) across frequency bins (x-axis) according to group (blue: GWIV, red: HGWV, yellow: CC). The mean power values across channels were not significantly different between groups in any frequency band in NREM sleep (ANOVA p > .05 in all frequency bins). In REM sleep, ANOVA revealed a significant difference among groups in the 33.33 – 36.17hz range, where the CC group had lower power than the GWI and HGWV groups (p < .05). CC = civilian control; GWIV = Gulf War illness veteran; HGWV = healthy Gulf War veteran

### Regional Reductions in NREM Sleep and REM Sleep EEG Power

After data was normalized to account for individual variations in absolute power by placing all individuals on the same scale (accomplished by z-scoring the absolute data across channels), groups were compared for topographic differences in EEG power distribution with suprathreshold cluster testing. Clusters with reduced EEG power emerged in circumscribed regions overlying the frontal lobe in both GWIV and HGWV relative to CC subjects. **Figure 3** panels A-C depicts the NREM topography of the CC group (A) and its comparison to GWIV (B) and HGWV (C) at each frequency band, with channels belonging to significant clusters appearing as white dots. In the SWA band during NREM sleep, significant clusters were identified with reduced power in the GWIV (N channels = 15, p = 0.04) and HGWV (N = 21, p = 0.037) groups relative to the CC group. In addition, the GWIV group had significant clusters with reduced power compared to the CC group in the Theta (N = 19, p = 0.038), Beta (N = 16, p = 0.041), and Gamma (N = 14, p = 0.021) bands. The HGWV group also had significant clusters with reduced power compared to the CC group, in the Theta (N = 23, p = 0.035), Sigma (N = 18, p = 0.053), and Beta (N = 29, p = 0.013) bands. In REM sleep (panels D-F), both the GWIV (E) and HGWV (F) groups had a general trend of lower power in frontal regions compared to the CC (D) group. However, significant clusters of channels only appeared in the GWIV group in the Sigma band (N = 19, p = 0.038), and the HGWV group in the SWA (N = 23, p = 0.015), Sigma (N = 29, p = 0.015), Beta (N = 37, p = 0.003), and gamma (N = 22, p = 0.024) bands. A direct topographic comparison between GWIV and GWV revealed a significant cluster of lower NREM gamma power in the GWIV group (N = 14, p = .049) (data not shown).

**Figure 3.**
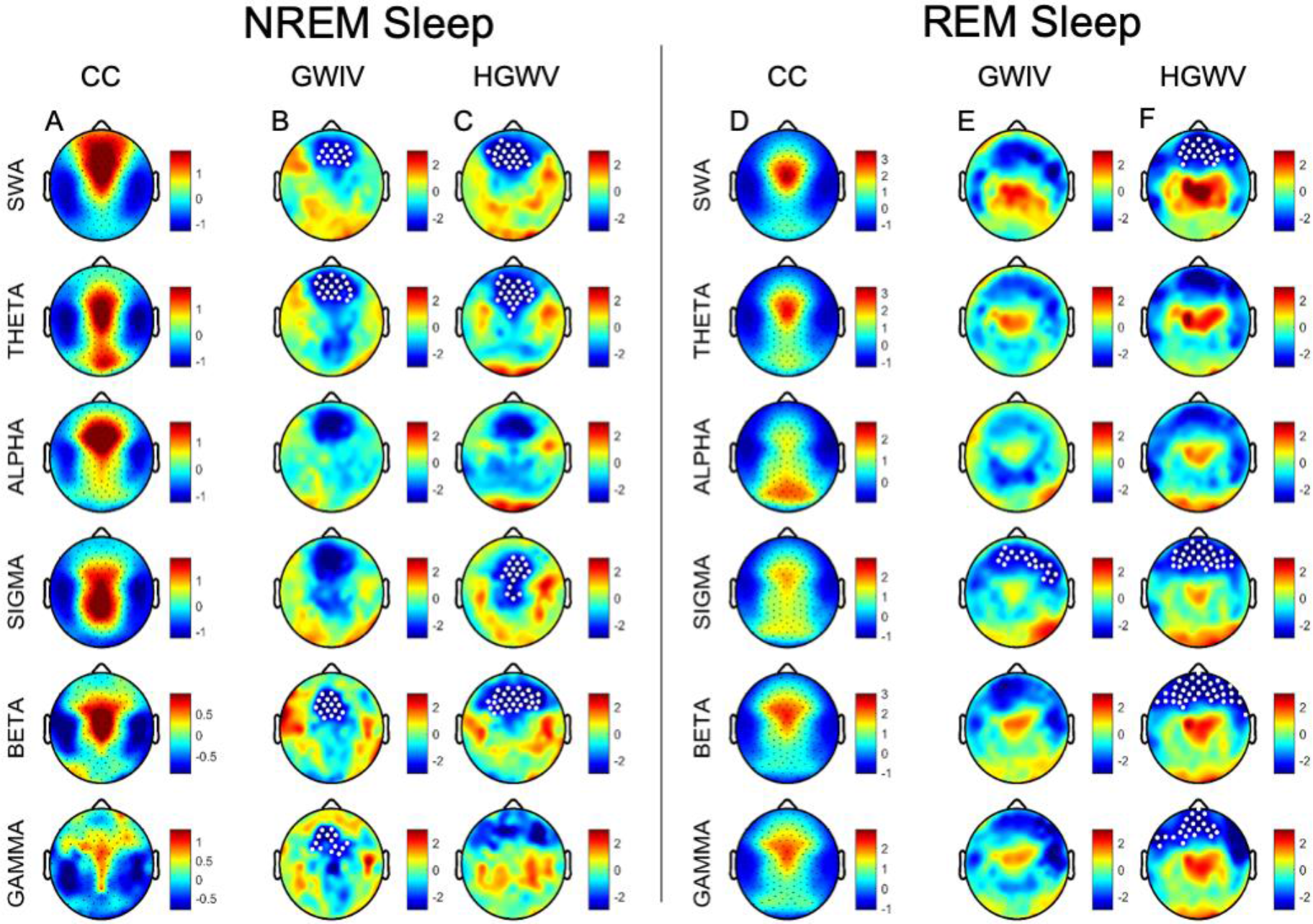
Topographical comparison reveals frontal clusters with reduced EEG power in both Gulf War veteran groups relative to CC in NREM and REM sleep. Panel A shows the normalized distribution of EEG power in the CC group in each frequency band (SWA: 1-4hz, Theta: 4-8hz, Alpha: 8-12hz, Sigma: 12-16hz, Beta:16-25hz, Gamma: 25-40hz). Panels B and C illustrate t-values of unpaired channel-to-channel t-tests of normalized EEG power between CC and GWIV (B), and between CC and HGWV (C) in each frequency band; darker blue represents more negative t-values and darker red more positive t-values. White dots represent significant clustering of channels with t-values exceeding a critical t-value, indicating significant regional differences between groups as identified by statistical nonparametric mapping with suprathreshold cluster testing. Similarly, REM sleep is illustrated at right, identically to above, however, CC, GWIV, and HGWV fall under panels D, E, and F, respectively. CC = civilian control; GWIV = Gulf War illness veteran; HGWV = healthy Gulf War veteran; REM = rapid eye movement sleep; SWA = slow wave activity

### Relationships with FSS and ISI

To assess the relationship between regional differences in sleep quality (as assessed by SWA) and symptom severity on the ISI and the FSS, we averaged z-scored power in the cluster of channels (N=23) that were significantly lower in the HGWV and GWIV groups relative to the CC group. Average SWA power for each individual was then separately correlated with scores on the FSS and ISI. Neither correlation was found to be significant (FSS: Pearson’s R = −0.02, p = 0.902; ISI: Pearson’s R = −0.33, p = .094).

## DISCUSSION

In this study, we characterize the distribution of HDEEG sleep power in male veterans who deployed in support of the 1990-1991 Persian Gulf War. Healthy veterans and veterans with fatigue-prominent GWI (i.e., HGWV and GWIV, respectively) were compared to age, sex, and OSA-comparable healthy controls (CC). Despite an absence of differences between groups in sleep macrostructure variables, topographic analysis of neural activity across the night revealed a broadband reduction of sleep EEG power in circumscribed regions overlying the frontal lobe for *both* GWIV *and* HGWV compared to CC. This frontal reduction in neural activity was present, to some extent, across all frequency bands during NREM sleep and REM sleep in all Persian Gulf War veterans. This regional reduction in frontal power overlying the frontal lobe was present in *both* GWIV *and* HGWV groups relative to the CC group, irrespective of fatigue and insomnia symptoms. Notably, psychiatric diagnoses do not explain our functional neurophysiological findings, as such clinical presentations represented study exclusion criteria.

Given the lack of exposure and deployment information for these veterans, assumptions regarding the causality of differences among groups are not possible. Deficits in SWA are widely reported to impair sleep’s restorative functions and are associated with self-reported poor sleep quality (39, 40). However, in our groups, there was no relationship between reduced frontal HDEEG power in SWA and either symptoms of fatigue or insomnia. Both fatigue prominent GWIV *and* asymptomatic HGWV displayed decreased regional sleep power relative to controls. These findings may point to subclinical neurodegenerative processes. Additionally, they may suggest that fatigue occurs independently of frontal lobe deficits for the GWIV group observed in this study. To be clear, interpretations of our findings remain speculative and would require validation. Taken together though, our results emphasize that a high level of clinical suspicion must be exercised during the evaluation of previously deployed veterans of the Persian Gulf War.

Our results fit with the literature wherein GWI veterans demonstrate frontal lobe dysfunction. Distorted executive function and response inhibition is reported within functional MRI and HDEEG-based event related potential studies (13, 14). GWI veterans display WM structural alterations and GM volume reduction within the frontal lobe (8-11). Cortical thinning, a measure of GM integrity, correlates with decreased slow wave density during sleep in healthy adults (41). Such cortical volume loss occurs in the setting of normal aging and has been shown to occur predominantly in the frontal regions (42, 43). In line with the findings herein, our lab has reported local broadband reductions of HDEEG sleep power (albeit over the parietal lobe) in OSA (30); these functional observations colocalize with MRI studies that show aberrations in parietal gray and white matter - findings thought to correlate with neuronal cell damage (44-47).

We do not possess radiologic imaging for veterans within this study, thus it remains unclear whether the HDEEG functional deficits observed herein correlate with structural changes in the frontal lobe. However, a number of studies have reported broad ranging CNS alterations including reduced gray matter volume (12, 15) as well as altered gray matter activity in response to behavioral, sensory and chemical stimuli (9, 10, 48-50). A recent study also identified microstructural changes in grey matter neurite density (NDI) in GWI veterans relative to their non-afflicted counterparts (51). However, grey matter NDI differences were less pronounced than all observed white matter abnormalities, including NDI as well as orientation dispersion, leading the authors to suggest that white matter abnormalities may represent a more robust marker for distinguishing veterans with GWI from control veterans. Either way, our observations add credence to the Institute of Medicine’s guidance in providing “follow-up assessments of Gulf War veterans for neurodegenerative diseases that have long latencies and are associated with aging” (1).

Gulf War veterans who deployed to Kuwait and Iraq encountered a litany of toxins while in theatre: pesticides, depleted uranium, sarin and cyclosarin gas, smoke from oil-well fires, petroleum-based combustion products, and prophylactic use of pyridostigmine bromide (1, 3). Mechanistic explanations for GWI focus upon the impact of these exposures on the cholinergic nervous system (4, 5). We do not have specific exposure histories to correlate among our veteran groups, and we did not specifically assess peripheral or central cholinergics. However, decreased tone of the peripheral parasympathetic nervous system is thought to contribute to fatigue in fibromyalgia and chronic fatigue syndrome (52). GWI has been paralleled to these two illnesses (1). Peripheral cholinergic dysfunction has been verified in GWI veterans, as represented by blunting of high frequency heart rate variability during sleep (4, 23). Fibromyalgia and chronic fatigue syndrome show similar dysautonomia (53-56). Interestingly, dysfunction in central cholinergic function has been implicated among veterans with GWI as well (5, 20). Thus, to postulate, peripheral cholinergic dysfunction might have contributed to fatigue symptoms in our study cohort. However, in contrast to our results, these studies’ veteran controls (Persian Gulf War veterans without GWI) did not display dysfunction. This discrepancy may point to the subtlety of presentations in Persian Gulf War veterans and/or reinforce the difficulty in comparing cohorts of Persian Gulf War veterans across studies when detailed deployment histories remain hard to come by. Nonetheless, the findings herein help to orient future directions.

The current results have both research and clinical implications. Additional HDEEG prospective research could aim to verify our findings. Especially interesting would be the correlation of functional HDEEG findings with intrasubject structural imaging to assess for WM or GM changes or volume loss. Further studies could correlate frontal lobe performance tasks and autonomic nervous system activity with functional and structural data. Such studies may help to elucidate underlying pathophysiology and thus determine treatment avenues. Clinically, our findings, and findings that may arise from subsequent research, could assist clinicians in identifying neurological, sleep, or other deficits in veterans of the Persian Gulf war. This, especially as the Persian Gulf War veteran population continues to age.

This study has strengths and weaknesses. The use of HDEEG represents a strength, as it allows for rigorous analysis of regional neural activity that are not possible with standard (12 channel) EEG montages. Overall, study participants were well-matched between groups and devoid of pharmaceuticals and comorbidities that impact sleep physiology. Small sample size is a weakness, which may cause overestimation of effects and thus false-positive results. We aimed to mitigate this by utilization of non-parametric statistical methods. This approach makes limited assumptions about data distribution and proves robust to outliers. Moreover, our results were not single-subject driven, as participant variability was evenly distributed. We lack corroborating radiographic imaging, clinical histories, cognitive or behavioral assessments, and long-term follow-up. Lastly, we were unable to attain information regarding veterans’ toxin exposures, deployment dates and geographic locations of operations. Yet, all veterans were recruited from the Persian Gulf War Registry, which is open only to those who deployed to Iraq or Kuwait for Operation Desert Storm (57).

## CONCLUSION

In sum, for cohorts of Persian Gulf War veterans - irrespective of GWI diagnoses and subjective fatigue - we observed broadband frontal EEG power reductions during sleep. It remains unclear if these findings correlate with structural or task-specific frontal lobe deficits. Our findings suggest that fatigue in GWI occurs independent of decreased sleep power. Future studies could aim to correlate sleep EEG power with frontal lobe performance tasks and structural integrity. Studies could also investigate how symptoms of fatigue in GWI correlate with peripheral nervous system function. Our findings highlight the importance of maintaining a high index of suspicion when caring for Persian Gulf War veterans in the clinical context.

## Data Availability

Data is available to interested researchers on request to

## Abbreviations

CC: civilian control
FSS: Fatigue Severity Scale
GM: gray matter
GWI: Gulf War illness
GWIV: Gulf War illness veteran
HDEEG: high-density EEG
HGWV: healthy Gulf War veteran
ISI: Insomnia Severity Index
NREM: non-rapid eye movement
OSA: obstructive sleep apnea
REM: rapid eye movement
WM: white matter

## Author Contributions

EM: formal analysis, writing (original draft; review & editing). SJ: conceptualization, formal analysis, project administration, supervision, writing (review & editing). BR: conceptualization, formal analysis, methodology, writing (review & editing). TS: investigation, data collection. TJ: funding acquisition, project administration, writing (review & editing).

## Acknowledgements

This work was supported by Department of Defense grant number W81XWH-10-2-0129. “Homeostatic and Circadian Abnormalities in Sleep and Arousal in Gulf War Syndrome.” Principal Investigator: Timothy M. Juergens, MD.

